# The 2022 RSV surge was driven by multiple viral lineages

**DOI:** 10.1101/2023.01.04.23284195

**Authors:** Gordon Adams, Gage K. Moreno, Brittany A. Petros, Rockib Uddin, Zoe Levine, Ben Kotzen, Katelyn Messer, Sabrina T. Dobbins, Katherine C. DeRuff, Christine Loreth, Taylor Brock-Fisher, Stephen F. Schaffner, Sushma Chaluvadi, Sanjat Kanjilal, Jeremy Luban, Al Ozonoff, Daniel Park, Sarah Turbett, Katherine J. Siddle, Bronwyn L. MacInnis, Pardis Sabeti, Jacob Lemieux

## Abstract

The US experienced an early and severe respiratory syncytial virus (RSV) surge in autumn 2022. Despite the pressure this has put on hospitals and care centers, the factors promoting the surge in cases are unknown. To investigate whether viral characteristics contributed to the extent or severity of the surge, we sequenced 105 RSV-positive specimens from symptomatic patients diagnosed with RSV who presented to the Massachusetts General Hospital (MGH) and its outpatient practices in the Greater Boston Area. Genomic analysis of the resulting 77 genomes (54 with >80% coverage, and 23 with >5% coverage) demonstrated that the surge was driven by multiple lineages of RSV-A (91%; 70/77) and RSV-B (9%; 7/77). Phylogenetic analysis of all US RSV-A revealed 12 clades, 4 of which contained Massachusetts and Washington genomes. These clades individually had times to most recent common ancestor (tMRCA) between 2014 and 2017, and together had a tMRCA of 2009, suggesting that they emerged well before the COVID-19 pandemic. Similarly, the RSV-B genomes had a tMRCA between 2016 and 2019. We found that the RSV-A and RSV-B genomes in our sample did not differ statistically from the estimated clock rate of the larger phylogenetic tree (10.6 and 12.4 substitutions per year, respectively). In summary, the polyphyletic nature of viral genomes sequenced in the US during the autumn 2022 surge is inconsistent with the emergence of a single, highly transmissible causal RSV lineage.

## Main

The United States experienced an unusually early, severe surge in respiratory syncytial virus (RSV) disease in autumn 2022 (**Figure (Fig) 1A, B**)^1^ for which the causes are not known. To investigate whether the emergence of a highly transmissible or virulent variant contributed to the surge, we sequenced RSV genomes from symptomatic patients diagnosed with RSV infection presenting to the Massachusetts General Hospital (MGH) and its outpatient practices in the Greater Boston Area. This cohort was broadly representative, demographically and clinically, of national RSV patients (**Supplemental (S) Tables 1A-C; Fig S1**). Whole genome sequencing of 105 residual diagnostic upper respiratory tract specimens from this cohort produced 54 near-complete (>80% coverage) and 23 partial (>5% coverage) RSV genomes, primarily from samples with higher viral loads (cycle threshold < 30; **Fig S2**). The sequencing data also revealed viral respiratory co-infections including a rhinovirus or enterovirus (9/105) and metapneumovirus (1/105; **Fig S3**).

**Figure 1:**
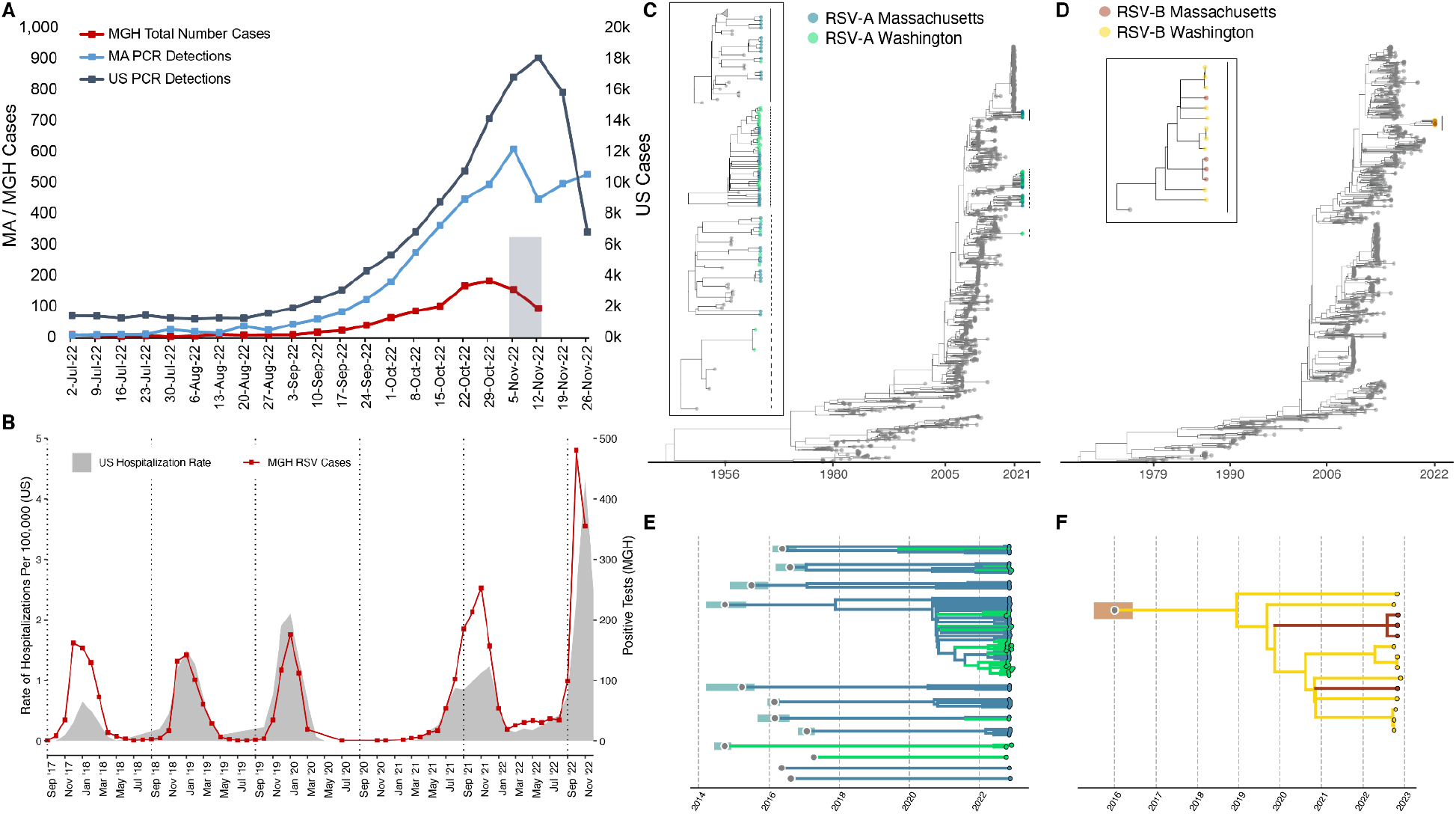
Epidemiological and genomic trends of the 2022 RSV surge. A) The number of PCR positive tests for RSV reported by the CDC in MA (blue, left axis) and the US (slate gray, right axis), along with the number of RSV positive tests conducted at MGH (red, left axis). The 105 sequenced samples were drawn from the gray Nov 2 - Nov 15 window. B) RSV hospitalization rates for CDC’s RSV-NET (shaded gray) and MGH RSV cases (red) for 2017 - 2022. (Pearson r = 0.82; p < 0.0001 via permutation). C) Maximum likelihood tree of all RSV-A genomes (N=1,267) (MA tips in blue, WA tips in green, others in gray). The tMRCA for 2022 RSV-A genomes was no later than 2008 (ML CI: 2008-04, 2008-09). In the box are zoomed in plots of the clades containing MA and WA with lines corresponding to clades on the tree. D) Maximum likelihood tree of all RSV-B genomes (N=944) (MA tips in orange, WA tips in yellow, others in gray). The tMCRCA for 2022 RSV-B genomes was approximately 2016 (ML CI: 2015-08, 2016-08). In the box are zoomed in plots of the clades containing MA and WA with lines corresponding to clades on the tree. Explosion plot of E) RSV-A and F) RSV-B lineages circulating in autumn 2022 with the inferred tMRCA (gray dots) and associated confidence intervals (shaded regions) for each lineage.

Genomic analysis demonstrated that the surge was driven by multiple lineages of RSV-A (91%; 70/77) and RSV-B (9%; 7/77) (**Fig 1C**,**D; Fig S4A**,**B**). All near-complete genomes were genotype GA2.3.5 (RSV-A) or GB5.0.5a (RSV-B). RSV-A genomes belonged to at least 10 distinct lineages, each with a time of their most recent common ancestor (tMRCA) between 2014 and 2017 (**Fig 1C**,**E; Fig S4A; Fig S5)**. The 4 complete RSV-B genomes similarly yielded a tMRCA estimate of 2019 as the later bound (**Fig 1D**,**F; Fig S4B; Fig S5**). Currently, the only other RSV genomes from the 2022 surge in the US are from Washington^2^ (WA; N=39); the RSV-A genomes from WA belong to 6 lineages, of which 4 also contain MA genomes (**Fig 1C, E; Fig S4A**,**B**). The genetic divergence of the 2022 RSV-A and RSV-B genomes was consistent with our estimated clock rate from the larger phylogenetic tree (10.6 and 12.4 substitutions per year, respectively), in contrast with the accelerated evolution seen with the highly transmissible SARS-CoV-2 Alpha, Delta, and Omicron variants^3–5^.

In summary, the 2022 RSV surge in the United States consisted of numerous pre-existing viral lineages, many shared between geographically disparate areas, a finding that is inconsistent with the emergence of a single, highly transmissible RSV lineage as the cause of the surge. The influence of non-viral factors on RSV transmission and virulences—including changes in immunity due to altered and suppressed RSV dynamics during the COVID-19 pandemic—should be investigated.

## Data Availability

Raw reads and assembled genomes are submitted to Genbank under PRJNA904288. Files associated with the Bayesian analyses are available on GitHub at https://github.com/bpetros95/rsv-2022.

## Acknowledgements

This work was sponsored in part by the Centers for Disease Control Broad Agency Announcement (75D30120C09605 to B.L.M.), the Howard Hughes Medical Institute Investigator award to P.C.S., the National Institute of Allergy and Infectious Diseases (U19AI110818 to P.C.S), the Massachusetts Consortium on Pathogen Readiness (to J.E.L and to J.L.), and the National Institute of General Medical Sciences (T32GM007753 and T32GM144273 to B.A.P. and Z.C.L.). The content is solely the responsibility of the authors and does not necessarily represent the official views of the National Institute of General Medical Sciences or the National Institutes of Health.

## Use of data

Raw reads and assembled genomes are submitted to Genbank under PRJNA904288. The data are available immediately and shared under Genbank’s use agreements to facilitate accelerated public health and scientific investigations. Files associated with the Bayesian analyses are available on GitHub at https://github.com/bpetros95/rsv-2022.

## Ethical statement

This study was conducted at the Broad Institute and Massachusetts General Hospital with approval from the Massachusetts General Brigham Institutional Review Board under Protocol #2019P003305 and from the MIT Institutional Review Board under Protocol #1612793224.

