## Supplemental Material for "The 2022 RSV surge was driven by multiple viral lineages"

**Description of the cohort:**

In autumn 2022, MGH reported a surge in RSV cases, with a total of 950 RSV-positive samples from Sept 1 – Nov 17 (**Supplemental Table 1**). In the MGH dataset, the number of positive RSV cases peaked the week of October 29, 2022, much earlier than previous years, and subsequently fell, mirroring trends that occurred at the state (**Figure 1A**) and national level (Pearson's correlation coefficient = 0.82, $p < 0.0001$; **Figure 1AB**)[1], an anomaly also noted globally[2].

The 77 patients from whom near-complete and partial genomes were assembled ("sequenced cases") consisted of 44% (n = 34) males with a median age of 2 years, and with 18% (n = 14) collected in inpatient care, 43% (n = 33) collected in outpatient care, and 39% (n = 30) collected in the emergency department (**Supplemental Table 1B**). All 77 patients reported symptoms consistent with a respiratory infection (**Supplemental Table 1B**). Sequenced cases were representative of the 950 patients in the greater MGH RSV-positive patient pool with respect to multiple demographic criteria, including sex (p = 0.12, chi-square test), age (p = 0.25, Wilcoxon rank sum test), presenting hospital unit (p = 0.07, chi-square test), and presence of symptoms (p = 1, chi-square test). (**Supplemental Table 1A and B**). The MGH cohorts are also consistent with the national RSV hospitalization data[1] with respect to sex ($p > 0.05$, chi-square tests; **Supplemental Table 1C**) though they are slightly older than the national hospitalized cohort (median age 2 at MGH vs. < 1 nationally, $p <0.0001$, **Supplemental Figure 1**).

We identified ten instances of co-infection with RSV and another known respiratory virus (9/105 rhinovirus or enterovirus, 1/105 metapneumovirus virus). We did not detect the presence of other common respiratory viruses (**Supplemental Figure 3**). We identified a single pair of near-complete genomes identical at the consensus-level, suggestive of a transmission link. Otherwise, genomic distances between samples ranged from 2 to 273 mutations for RSV-A, and 3 to 70 mutations for RSV-B, with a mean pairwise distance of 181 and 36 mutations respectively.

This cohort also reflects the MGH outpatient testing recommendations for RSV, where the the Xpert Xpress SARS-CoV-2/Flu/RSV (Cepheid, Sunnyvale, CA) panel was recommended for symptomatic children under age 3, immunocompromised patients, and individuals with chronic lung disease.

**Methods**

***Sample collection***

We obtained 105 frozen (-80°C), archived, de-identified upper respiratory tract samples that were positive for RSV from the MGH Clinical Microbiology Laboratory. Samples were from symptomatic individuals presenting for clinical care at MGH and its affiliated outpatient practices, and were selected to deeply sample the epidemic over two November epi-weeks (Nov 2 – Nov 15). Samples were tested for the presence of viral respiratory pathogens by one of three clinically verified, Food and Drug Administration (FDA) emergency use authorized or approved assays: the Xpert Xpress SARS-CoV-2/Flu/RSV (Cepheid, Sunnyvale, CA; 90/105 samples) or the BioFire Respiratory 2.1 Panel (BioFire Diagnostics, Salt Lake City, UT; 15/105 samples). The Xpert Xpress SARS-CoV-2/FLU/RSV assays contain targets for the detection of SARS-CoV-2, influenza A and B, and RSV. The BioFire Respiratory 2.1 Panel contains targets for the following viruses: SARS-CoV-2, non-SARS-CoV-2 coronaviruses, human rhinovirus/enterovirus (combined target), influenza A and B, RSV, parainfluenza viruses 1-4, adenovirus, and human metapneumovirus.

#### *Processing and analysis of national data*

Weekly 2022 national case counts were downloaded from the Centers for Disease Control and Prevention[3], and weekly national hospitalization rates (with demographic breakdowns by age and sex) were downloaded from the RSV Hospitalization Surveillance Network (RSV-NET)[1]. United States Census data[4] were used to convert hospitalization rates per 100,000 individuals of a particular demographic status into approximate case counts.

Monthly national case counts were compared with MGH's monthly positive test counts. Months with missing data in either data set were removed, and the Pearson correlation was calculated. To determine the significance of this correlation, the monthly national case counts were then scrambled across dates 10,000 times, generating a null distribution of correlation coefficients (**Figure 1B**).

The national hospitalization data binned ages (0-6 months, 6-12 months, 1-2 years, 2-5 years, 5-11 years, 12-17 years, 18-29 years, 30-39 years, 40-49 years, 50-64 years, 65-74 years, 75-84 years, 85+ years). For each case in each age bin, we randomly sampled an age uniformly across the bin. These sampled ages were compared to the ages of individuals in the MGH cohorts using the Wilcoxon rank sum test (**Supplemental Figure 1**).

#### *RSV quantification and sequencing*

To prepare samples for sequencing, we extracted total nucleic acid from upper respiratory tract samples in transport media, removed DNA with DNase I treatment, and assessed viral quantity using an RSV-specific SYBR green RT-qPCR assay[5]. Human ribosomal RNA was then depleted with an RNase-H based method, and libraries were constructed using a strand-specific ligation-based approach[6]. Briefly, RNA was heat-fragmented and first-strand cDNA was generated with randomly primed reverse transcription. Second-strand cDNA was generated with nick translation and labeled with dUTP. Full-length, y-shaped, unique-dual-index Illumina sequencing adapters containing a UMI adjacent to the i7 index were ligated to the resulting double-stranded DNA. Samples were then treated with Uracil-Specific Excision Reagent (USER enzyme) to excise dUTP from the second strand. Libraries were PCR amplified, quantified, and pooled in equimolar ratios for sequencing on Illumina MiSeq or NextSeq 550 instruments.

#### *RSV genome assembly and analysis*

To assemble RSV genomes, we conducted all analyses using viral-ngs 2.1.28[7] on the Terra platform (app.terra.bio). All of the workflows named below are publicly available via the Dockstore Tool Registry Service (dockstore.org/organizations/BroadInstitute/collections/pgs). Briefly, samples were demultiplexed, reads were filtered for known sequencing contaminants, and de novo assembly with scaffolding against RSV-A (GenBank: KY654518.1) and RSV-B (GenBank: MZ516105.1) was performed.

The mean unambiguous genome length of these genomes was 11,970 bp for RSV-A and 10,268 bases for RSV-B. Assembled genomes with ≥50% completeness were deposited into NCBI GenBank. Raw reads for all samples (including those that did not produce a successful genome) were deposited in NCBI SRA. All NCBI data were deposited under BioProject PRJNA904288.

#### *Phylogenetic analysis*

We constructed RSV-A and RSV-B specific maximum-likelihood (ML) phylogenetic trees[8] with associated visualizations using an augur pipeline[9] (*augur_from_assemblies*), part of the Nextstrain project[10]. We included all contextual genomes from GenBank with reported collection dates and a genome length greater than 12,160 bp, corresponding to 80% of the reference genome length (resulting in a total of RSV-A N=1,238; RSV-B N=934 genomes downloaded on December 6, 2022). Within augur, RSV-A and RSV-B genomes were aligned via MAFFT v.7 to GenBank NC_038235.1 and NC_001781.1, respectively. This alignment was further processed in both the ML and Bayesian analyses.

We used the contextualized ML phylogeny to calculate the number of lineages circulating and estimate the time to tMRCA. To do so, we assigned a binary trait to each genome in the phylogeny, associated with the genome division of collection, and used Nextstrain's ancestral inference to infer the state of that trait for each internal node in the tree. We defined a lineage at the first node attributed to contain only MA descendants. Using baltic[11], we extracted these changes from the phylogenetic tree and plotted the inferred tMRCA for each lineage using matplotlib[12].

In parallel, we conducted molecular dating using BEAST version 1.10.5[13]. We used the same genome length filter (80% of the reference genome) and alignment described in the ML analysis. However, we used a subset of available genomes, selected as follows: (i) all genomes generated in 2022 (79 for RSV-A, 14 for RSV-B); (ii) all descendants of the parent node of all 2022 genomes (4 additional genomes for RSV-A, 0 additional genomes for RSV-B); and (iii) genomes sampled uniformly across time (221 additional genomes for RSV-A, 290 additional genomes for RSV-B), to reach a total of 304 genomes each for RSV-A and RSV-B. In BEAUti v.1.10.4, we defined two taxon sets for which we generated posterior distributions of the tMRCAs: one including all 2022 (i.e., MA and WA) genomes and one containing solely the 2022 MA genomes created in this study. We used dates with variable precision (i.e., retained sequences with missing day or month resolution) and used the HKY substitution model[14] with 4 categories of gamma site heterogeneity[15]. We combined a strict clock model with a coalescent tree prior (a piecewise Bayesian skyline model[16] with 5 groups). We ran BEAST twice, each with a UPGMA starting tree and 300 million MCMC steps, sampled every 1000 steps. We removed the first 10% of steps as burn-in and combined the two .log files via LogCombiner v1.10.4. We analyzed results in Tracer v.1.7.2[17] to ensure convergence, and isolated the tMRCA estimates from the combined .log file. We also combined the two .trees files in LogCombiner v1.10.4, removing the first 10% as burn-in and sampling every 10,000 trees. We generated the maximum clade credibility (MCC) tree with median heights using TreeAnnotator v1.10.4[18]and displayed it using FigTree v1.4.4.

Using the molecular clock rate regression line calculated by augur, we determined the residuals (number of mutations) per sample. Samples with a residual falling outside of the 2.5–97.5th percentiles were considered to statistically deviate from the molecular clock rate.

#### *Coinfection analysis*

One of the 15 samples tested by BioFire Respiratory 2.1 Panel was positive for rhinovirus/enterovirus, prompting interest in coinfections. To assess for additional coinfections in all samples that underwent unbiased sequencing, taxonomic classification of reads was performed with Kraken2[19] using a custom database as previously described[20]. A viral taxon was considered present if >10 reads were assigned to it. We filtered the results to include solely known human respiratory viruses. All Kraken2 classifications were verified by megablast as follows: first, megablast was run on *de novo* contigs. If the least common ancestor of the top e-value megablast hit agreed with the Kraken2 classification for at least one contig (≥200 bp), we considered the taxon present. If this criterion was not met, megablast was run on all reads assigned to the taxon of interest by Kraken2. If the least common ancestor of the top e-value megablast hits agreed with the Kraken2 classification for at least 90% of the assigned reads, the taxon was considered present

### Figures

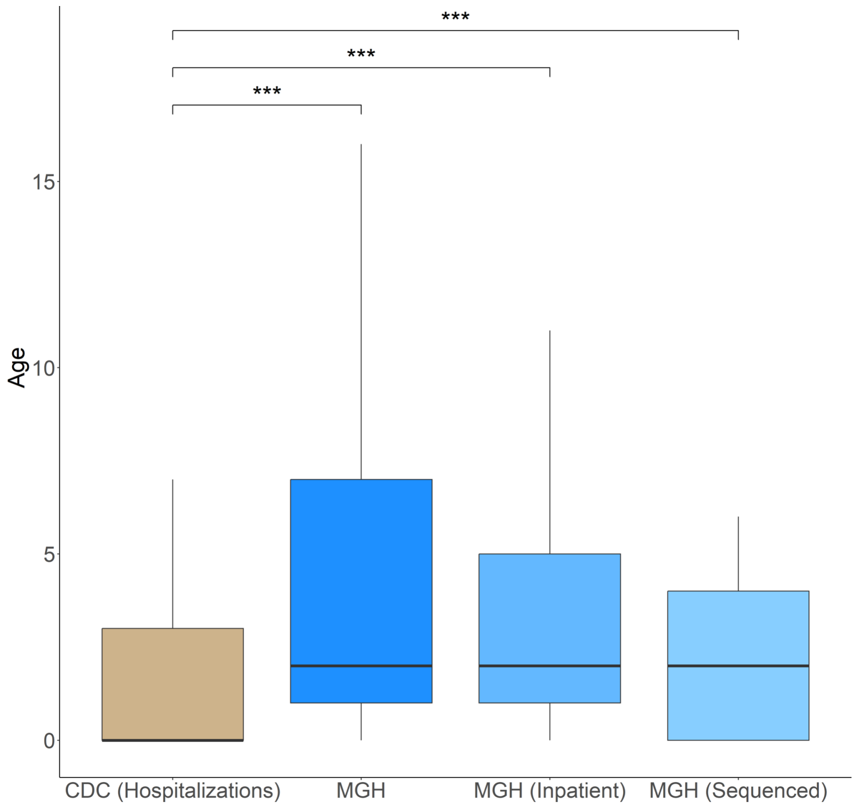


**Supplemental Figure 1: Age of individuals in the national and MGH data sets.** Median ages, < 1 year (CDC hospitalization data), 2 (MGH patients, N = 950), 2 (MGH hospitalized patients, N = 212), and 2 (MGH sequenced cohort, N = 77). ***, $p < 0.0001$ via Wilcoxon rank sum test.

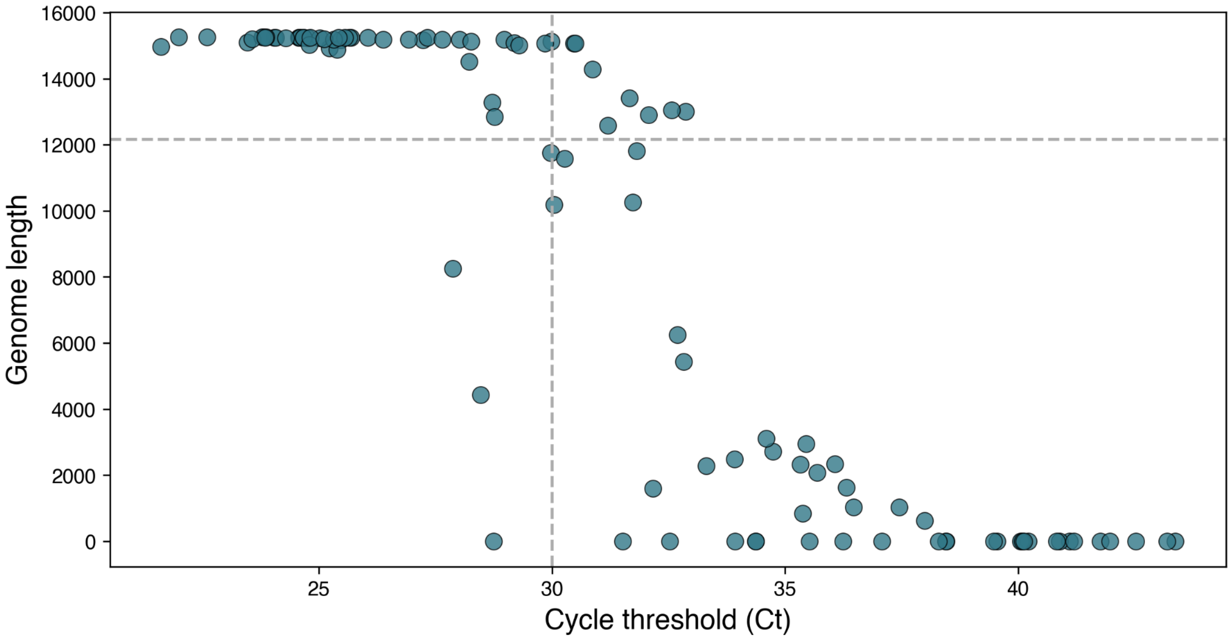


**Supplemental Figure 2: Unambiguous genome length vs. cycle threshold (Ct).** Each dot represents a unique sample in our dataset. Dashed lines are at a Ct of 30, and an unambiguous genome length of 12160, corresponding to ~80% of the RSV genome. Of the 105 samples, 92% (46/50) with a RT-qPCR Ct value ≤30 resulted in a genome, whereas only 15% (8/55) with a Ct >30 resulted in a genome (Fisher's exact test, p<0.001).

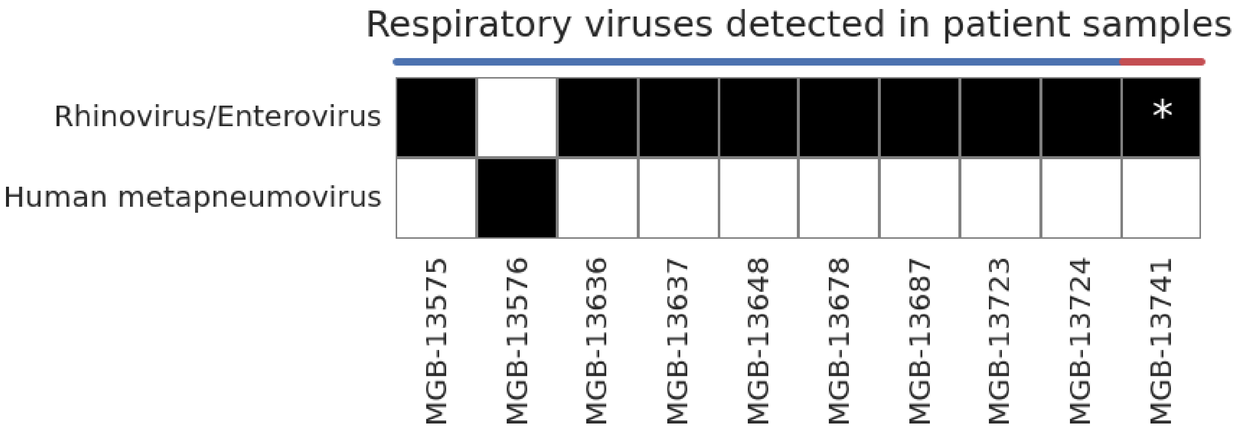


**Supplemental Figure 3: Coinfection analysis of unbiased sequencing reads.** Respiratory viral co-infections detected in patient samples by Kraken2 and megablast (Methods). Line indicates clinical testing with Xpert Xpress SARS-CoV-2/Flu/RSV (blue) or BioFire Respiratory 2.1 Panel (red). Star (*) indicates a positive clinical test.

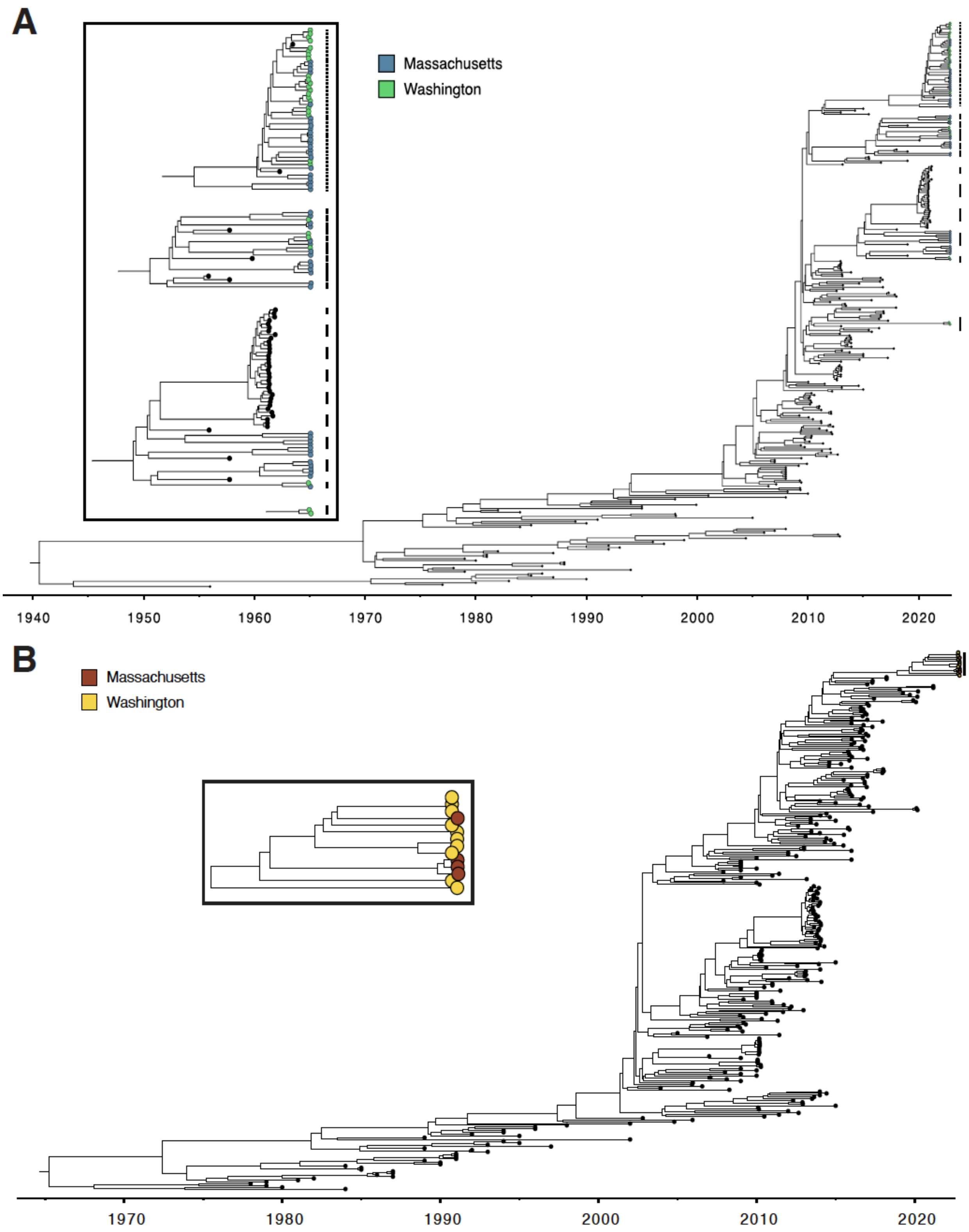
A
Massachusetts
Washington
1940
1950
1960
1970
1980
1990
2000
2010
2020
B
Massachusetts
Washington
1970
1980
1990
2000
2010
2020

**Supplemental Figure 4: Bayesian phylogenetic trees.** A) Phylogenetic tree of a subset of RSV-A genomes (N=304; MA tips in blue, WA tips in green, others in black). The inset displays clades containing 2022 genomes with lines denoting their locations in the larger tree. B) Phylogenetic tree of a subset of RSV-B genomes (N=304; MA tips in red, WA tips in yellow, others in black). The inset displays the clade containing 2022 genomes with a line denoting its location in the larger tree.

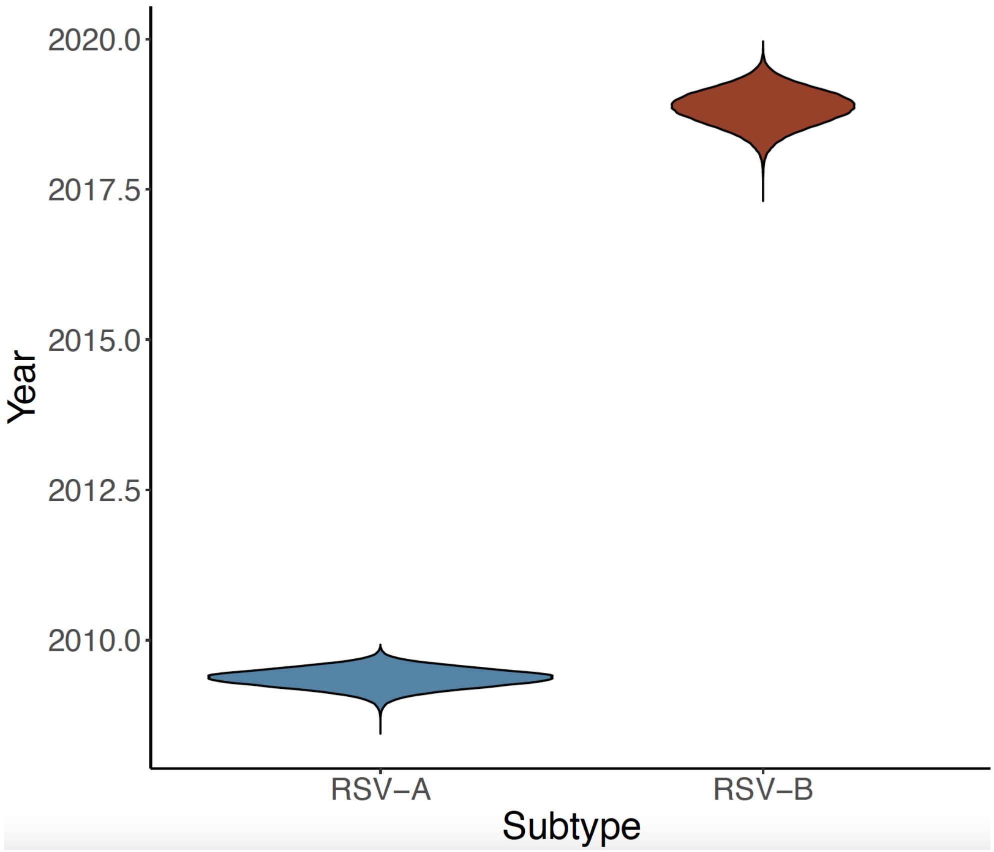


**Supplemental Figure 5: Molecular dating of the 2022 RSV samples.** A) Posterior distribution of the tMRCA for RSV-A (95% highest posterior density interval in blue, 2009-02 – 2009-08). B) Posterior distribution of the tMRCA for RSV-B (95% highest posterior density interval in red, 2018-03 – 2019-05).

**Supplemental Table 1A. MGH sample demographics.**

| | All Patients (N = 950) | Inpatient (N = 212) | Outpatient (N = 286) | Emergency Room (N = 452) |
|---|---|---|---|---|
| **Baseline Demographics** | | | | |
| Median age (IQR) - yr | 2 (0-5) | 2 (1-5) | 2 (0-6) | 2 (0-4.3) |
| Male - no. (%) | 507 (53.4) | 100 (47.2) | 162 (56.6) | 244 (54.0) |
| **Reported Symptoms** | | | | |
| Symptomatic - no. (%) | 950 (100) | 212 (100) | 286 (100) | 452 (100) |
| Cough - no. (%) | 719 (75.7) | 168 (79.2) | 229 (80.1) | 322 (71.2) |
| Fever - no. (%) | 507 (53.4) | 135 (63.7) | 142 (49.7) | 230 (50.9) |
| Sore Throat - no. (%) | 90 (9.5) | 21 (9.9) | 23 (8.0) | 46 (10.2) |
| Nasal Congestion/Runny Nose - no. (%) | 361 (38.0) | 92 (43.4) | 149 (52.1) | 120 (26.5) |
| Shortness of Breath - no. (%) | 199 (20.9) | 24 (11.3) | 24 (8.4) | 151 (33.4) |
| Loss of Smell or Taste - no. (%) | 4 (0.4) | 0 (0) | 1 (0.3) | 3 (0.7) |
| Muscle Aches - no. (%) | 28 (2.9) | 7 (3.3) | 13 (4.5) | 9 (2.0) |

**Supplemental Table 1B. Sequenced cohort demographics.**

| | All Patients (N = 77) | Inpatient (N = 14) | Outpatient (N = 33) | ER (N = 30) |
|---|---|---|---|---|
| **Baseline Demographics** | | | | |
| Median age (IQR) - yr | 2 (0-4) | 1.5 (0.3-5.5) | 1 (0-3) | 2 (0.3-15.5) |
| Male - no. (%) | 34 (44.2) | 5 (35.7) | 15 (45.5) | 14 (46.7) |
| **Reported Symptoms** | | | | |
| Symptomatic - no. (%) | 77 (100) | 14 (100) | 33 (100) | 30 (100) |
| Cough - no. (%) | 62 (80.5) | 9 (64.3) | 28 (84.8) | 25 (83.3) |
| Fever - no. (%) | 43 (55.8) | 9 (64.3) | 17 (51.5) | 17 (56.7) |
| Sore Throat - no. (%) | 11 (14.3) | 1 (7.1) | 3 (9.1) | 7 (23.3) |
| Nasal Congestion/Runny Nose - no. (%) | 37 (48.1) | 9 (64.3) | 19 (57.6) | 9 (30.0) |
| Shortness of Breath - no. (%) | 15 (19.5) | 0 (0) | 4 (12.1) | 11 (36.7) |
| Loss of Smell or Taste - no. (%) | 0 (0) | 0 (0) | 0 (0) | 0 (0) |
| Muscle Aches - no. (%) | 3 (3.9) | 0 (0) | 2 (6.1) | 1 (3.3) |
| **RSV Strain** | | | | |
| RSV-A - no. (%) | 70 (90.9) | 13 (92.9) | 28 (84.8) | 29 (96.7) |
| RSV-B - no. (%) | 7 (9.1) | 1 (7.1) | 5 (15.2) | 1 (3.3) |

**Supplemental Table 1C. Number of cases (MGH and sequenced cohorts) or hospitalization rate per 100,000 (CDC) by sex. P-values calculated relative to the CDC data using the chi-square test.**

| | Female | Male | P-Value |
|---|---|---|---|
| CDC (Hospitalizations per 100,000) | 24.8 | 27.1 | ----- |
| MGH | 444 | 506 | 0.09 |
| MGH (Inpatient) | 112 | 100 | 0.78 |
| MGH (Sequenced) | 43 | 34 | 0.44 |

15. Yang Z. Maximum likelihood phylogenetic estimation from DNA sequences with variable

rates over sites: approximate methods. J Mol Evol 1994;39(3):306–14.
